# A rare non-coding enhancer variant in *SCN5A* contributes to the high prevalence of Brugada syndrome in Thailand

**DOI:** 10.1101/2023.12.19.23299785

**Authors:** Roddy Walsh, John Mauleekoonphairoj, Isabella Mengarelli, Arie O. Verkerk, Fernanda M. Bosada, Karel van Duijvenboden, Yong Poovorawan, Wanwarang Wongcharoen, Boosamas Sutjaporn, Pharawee Wandee, Nitinan Chimparlee, Ronpichai Chokesuwattanaskul, Kornkiat Vongpaisarnsin, Piyawan Dangkao, Cheng-I Wu, Rafik Tadros, Ahmad S. Amin, Krystien V.V. Lieve, Pieter G. Postema, Maarten Kooyman, Leander Beekman, Dujdao Sahasatas, Montawatt Amnueypol, Rungroj Krittayaphong, Somchai Prechawat, Alisara Anannab, Pattarapong Makarawate, Tachapong Ngarmukos, Keerapa Phusanti, Gumpanart Veerakul, Zoya Kingsbury, Taksina Newington, Uma Maheswari, Mark T. Ross, Andrew Grace, Pier D. Lambiase, Elijah R. Behr, Jean-Jacques Schott, Richard Redon, Julien Barc, Vincent M. Christoffels, Arthur A.M. Wilde, Koonlawee Nademanee, Connie R. Bezzina, Apichai Khongphatthanayothin

## Abstract

Brugada syndrome (BrS) is a cardiac arrhythmia disorder that causes sudden death in young adults. Rare genetic variants in the *SCN5A* gene, encoding the Na_v_1.5 sodium channel, and common non-coding variants at this locus, are robustly associated with the condition. BrS is particularly prevalent in Southeast Asia but the underlying ancestry-specific factors remain largely unknown. Here, we performed genome sequencing of BrS probands from Thailand and population-matched controls and identified a rare non-coding variant in an *SCN5A* intronic enhancer that is highly enriched in BrS cases (3.9% in cases, odds ratio 20.2-45.2) and predicted to disrupt a Mef2 transcription factor binding site. Heterozygous introduction of the enhancer variant in human induced pluripotent stem cell-derived cardiomyocytes (hiPSC-CMs) caused significantly reduced *SCN5A* expression from the variant-containing allele and a 30% reduction in Na_v_1.5-mediated sodium-current density compared to isogenic controls. This is the first example of a validated rare non-coding variant at the *SCN5A* locus and partly explains the increased prevalence of BrS in this geographic region.

## Main

BrS is a cardiac arrhythmia disorder associated with increased risk of sudden death in young adults and characterised by ST-segment elevation in the right precordial leads of the electrocardiogram (ECG) occurring either spontaneously or after challenge with sodium channel blockers like ajmaline^1^. Rare loss-of-function coding variants in *SCN5A*, encoding the Na_v_1.5 sodium channel underlying the cardiac sodium current (I_Na_), are found in approximately 20% of European-ancestry BrS cases^2^. Several other genes have been implicated in BrS but were considered to have limited evidence of association by ClinGen re-evaluation and *SCN5A* remains the only clinically actionable gene^3,4^. Genome-wide association studies (GWAS) have demonstrated a strong contribution for common genetic variation to BrS susceptibility (SNP-based heritability, h2_SNP_, between 0.17-0.34), indicating the genetic architecture is more complex than a simple monogenic disease model^5^.

Eight of the 21 independent genome-wide significance signals detected by the BrS GWAS were located at the *SCN5A-SCN10A* locus, further cementing the central role for this genomic region in BrS genetic aetiology^5^. All lead variants are non-coding and located in or near regulatory elements (REs) at the *SCN5A-SCN10A* locus that have been characterised in detail (Fig. 1A), including the *SCN5A* promoter, a downstream super enhancer encompassing RE6-9 and two intragenic *SCN5A* enhancer elements (RE4/RE5)^6,7^. A short transcript of *SCN10A*, comprising the last 7 exons and controlled by an intronic enhancer-(RE1-2)-promoter complex, is expressed in the heart^8^. A common variant in RE1 associated with BrS and ECG traits, rs6801957, leads to reduced expression of *SCN10A-short* and decreased cardiac I_Na_ while minimally affecting *SCN5A* expression^8^.

**Fig. 1.**
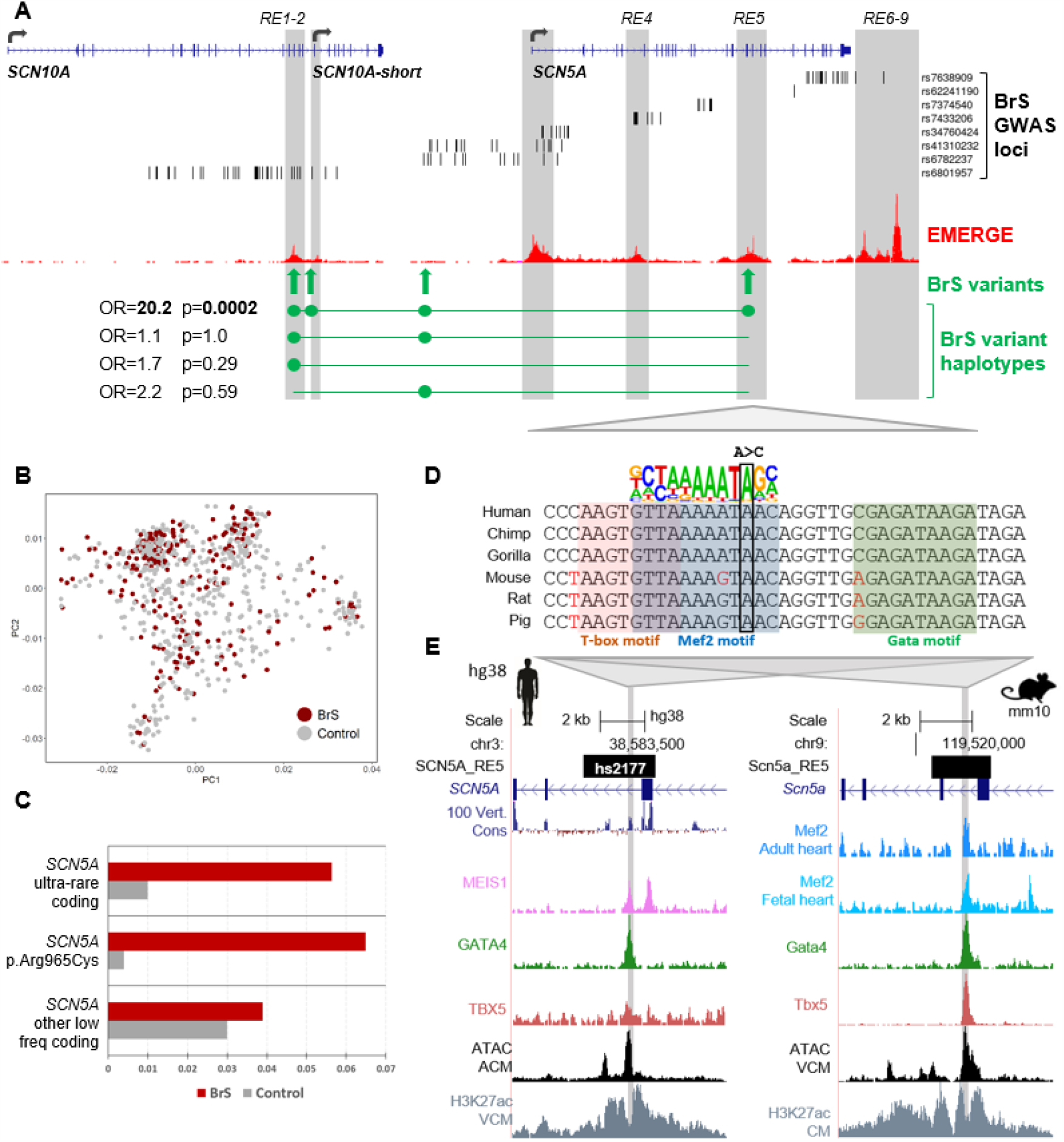
A) Overview of the coding sequence and regulatory elements at the SCN10A-SCN5A locus, highlighting SNPs from Brugada syndrome (BrS) genome-wide association study (lead SNP and SNPs with r^2^>0.5 in European populations), the EMERGE track of cardiac-specific epigenetic markers, the location of the four rare and low frequency non-coding variants associated with BrS in Thai patients and odds ratios (OR) and p values for each haplotype. B) Principal component analysis (PCA) for BrS cases and controls from Thailand. C) BrS case and control frequencies for SCN5A coding variants – ultra-rare variants (gnomAD FAF<0.00001), the low frequency variant p.Arg965Cys and other low frequency variants (gnomAD FAF<0.001). D) Sequence alignment of 6 mammalian species, incl. human and mouse, of the hs2177 enhancer core that encompasses the GRCh38:3-38580380-A-C variant (indicated by box). The variant is located in and disrupts a predicted and conserved Mef2 motif site (indicated by blue shading) and is flanked by T-box (red shading) and Gata (green shading) recognition sites as predicted by Homer. E) UCSC browser views of the hs2177-SCN5A RE5 enhancer in human (hg38) and mouse (mm10). Relevant cardiac transcription factor ChIP-seq and ATAC-seq traces from the literature were plotted (see Table S4 for dataset references). Abbreviations: ACM: atrial cardiomyocytes, VCM: ventricular cardiomyocytes, CM: cardiomyocytes.

BrS is several times more prevalent in East, and especially Southeast, Asia compared to regions of predominantly European ancestry. After its initial description 1992^9^, it was subsequently recognised as the major cause of sudden unexplained death syndrome in Thailand, a condition causing sudden nocturnal death in young men and traditionally locally known as “*Lai Tai*”^10,11^. The high rate of BrS in Southeast Asia is not due to an increased prevalence of ultra-rare loss-of-function *SCN5A* coding variants; indeed a relatively lower diagnostic yield is observed compared to European ancestry cases (∼5% vs 20%)^2,12^. We previously demonstrated enrichment of a low frequency *SCN5A* coding variant of intermediate effect size, p.Arg965Cys, in Thai BrS patients compared to population-matched controls^12^. However, other genetic factors underlying the higher prevalence of BrS in Southeast Asia are still largely unknown.

Here, we identify and functionally validate a novel rare non-coding variant in an *SCN5A* enhancer which is highly enriched in BrS patients from Thailand. Genome sequencing data was generated for BrS probands and controls from Thailand – see online methods for details of data generation, read mapping, variant calling and quality control (QC) checks. After sample-level QC, a total of 231 probands and 500 ancestry-matched controls were included, with principal component analysis demonstrating the cohorts were of comparable ancestral background (Fig. 1B). Of 231 BrS probands (mean age 47.6±12.8 years, 95% male), 88% had a documented spontaneous BrS ECG pattern and 76% were symptomatic (Table 1). 5.6% of cases had an ultra-rare (gnomAD filtering allele frequency (FAF)<0.00001) *SCN5A* coding variant (vs 1.0% in controls, p=4.0E-04), 6.5% carried the SCN5A:p.Arg965Cys low frequency variant (vs 0.4% in controls, p=1.6E-06) and 3.9% had other low frequency (gnomAD FAF<0.001) *SCN5A* coding variants, although the latter were only marginally enriched over controls (3.0%, p=0.51) (Fig. 1C, Table S1).

**Table 1:** Clinical characteristics of cases grouped according to SCN5A genotype (cases with low frequency SCN5A coding variants other than p.Arg965Cys were not separately evaluated). Clinical data is available for all 231 BrS cases, except for Spontaneous BrS ECG pattern (n=186 cases only), FH of SCA/SCD (n=192 cases only) and ECG parameters (case numbers as shown). ECG – electrocardiogram; FH – family history; SCA – sudden cardiac arrest; SCD – sudden cardiac death; VF – ventricular fibrillation; VT – ventricular tachycardia;

|  | All Cases<br>(n=231) | RE5<br>variant<br>(n=9) | Ultra-rare<br>coding variant<br>(n=13) | p.Arg965Cys<br>(n=15) | SCN5A variant<br>negative<br>(n=186) |
| --- | --- | --- | --- | --- | --- |
| Male sex, n (%) | 220 (95%) | 9 (100) | 13 (100) | 15 (100) | 175 (94%) |
| Age (mean ± SD) | 47.6±12.8 | 48.9±12.9 | 45.1±13.7 | 49.7±12.7 | 47.7±12.7 |
| Spontaneous BrS ECG pattern,<br>% (n) | 88 (164) | 75 (6) | 89 (8) | 100 (13) | 87 (130) |
| Symptomatic, % (n) | 75 (173) | 89 (8) | 77 (10) | 67 (10) | 76 (141) |
| SCA/VT/VF, % (n) | 58 (134) | 89 (8) | 54 (7) | 67 (10) | 58 (108) |
| Syncope, % (n) | 34 (78) | 44 (4) | 46 (6) | 27 (4) | 34 (63) |
| FH of SCA/SCD, % (n) | 42 (81) | 38 (3) | 60 (6) | 33 (5) | 43 (66) |
| **ECG parameters** |  |  |  |  |  |
| Cases with ECG data, n | 187 | 8 | 9 | 13 | 150 |
| RR interval, ms | 853±171 | 846±117 | 892±169 | 869±161 | 846±176 |
| PR interval, ms | 177±29 | 181±28 | 187±33 | 191±32 | 175±29 |
| QRS duration, ms | 109±19 | 111±25 | 110±18 | 111±12 | 109±20 |
| QT interval, ms | 432±44 | 434±39 | 409±27 | 438±33 | 433±46 |
| QRS axis (°) | 44±51 | 7.88±52 | 55.4±29 | 37.4±47 | 46.4±51 |
| Left QRS axis deviation, % (n) | 9(16) | 25(2) | 0(0) | 8(1) | 8(12) |

Rare and low frequency non-coding variants at the *SCN5A-SCN10A* locus encompassing the defined promoter and enhancer regions were assessed for enrichment in BrS probands versus controls. Four variants in linkage disequilibrium (LD, r^2^>0.2) were identified (Table 2) - chr3-38580380-A-C in the *SCN5A* enhancer RE5, chr3-38719550-C-T in the *SCN10A*-short transcript promoter, chr3-38724980-T-C in the *SCN10A*-short enhancer RE2 and chr3-38683338-C-T in the intergenic region between *SCN5A* and *SCN10A* (Fig. 1A) (GRCh38 coordinates).

**Table 2:**
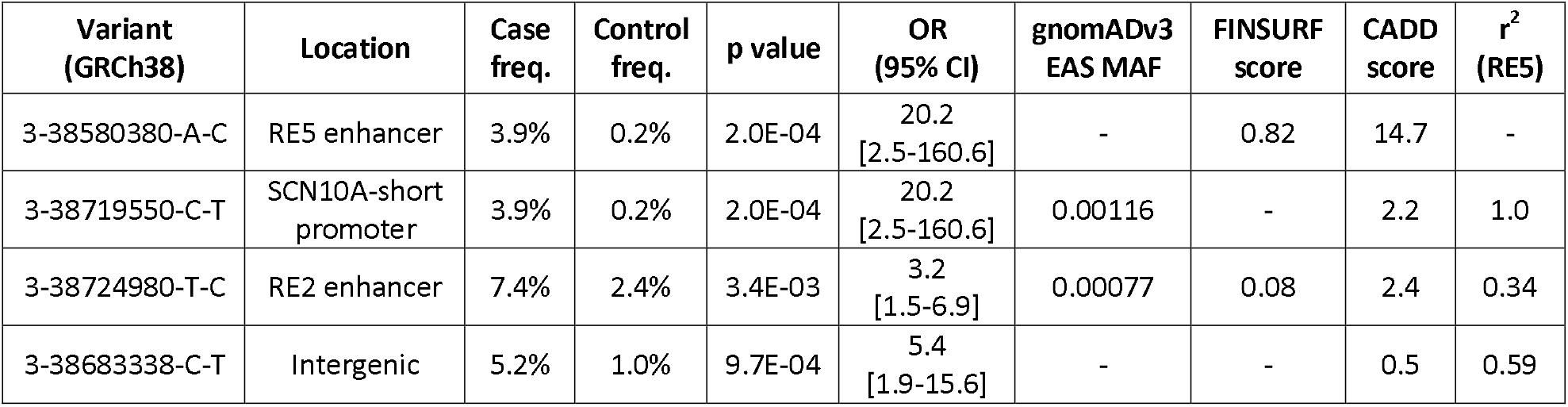
Details of the non-coding variants at the SCN5A-SCN10A locus associated with BrS in cases and controls from Thailand, including location with respect to SCN5A/SCN10A regulatory elements, sample-level frequencies in cases (n=231) and controls (n=500), Fisher’s exact test p values, odds ratios (OR) with 95% confidence intervals (CI), minor allele frequencies (MAF) in gnomADv3 East Asian (EAS) individuals, FINSURF score (a machine-learning approach to predict the functional impact of non-coding variants in regulatory regions, score ranges from 0 to 1 with increasing deleteriousness)^13^, CADD scores and r^2^ values of linkage disequilibrium with respect to the RE5 variant (calculated from the case and control data in this study).

For these four variants, the largest case-control odds ratios (OR) were observed for the RE5 and *SCN10A*-short promoter variants (in complete LD in this dataset). Haplotypes containing only one or both of the RE2 and intergenic variants had no association with BrS, confirming they are merely tagging the causal variant (Fig. 1A). The RE5 variant alters a nucleotide fully conserved across 35 mammalian species with available orthologous sequence data, in contrast to the other three variants (Fig. S1) and has higher computational deleteriousness scores from the CADD and FINSURF^13^ algorithms (Table 2). The RE5 variant is also absent in the gnomADv3 database (n=76,165 genomes including n=2,604 of East Asian (EAS) ancestry), whereas the *SCN10A*-short promoter variant has a gnomAD-EAS frequency of 0.001, indicating it also exists on haplotypes distinct from the RE5 variant. Based on this cumulative evidence, the RE5 variant was deemed to be the putative causal variant for this risk haplotype.

The RE5 variant was detected in 3.9% (n=9) of cases vs 0.2% (n=1) of controls (p=2.0e-04, OR=20.2 [2.5-160.6]). We searched the Genomics Thailand database, comprising 10,043 individuals with a range of clinical and pharmacogenetics indications (https://genomicsthailand.com/Genomic/home), to attain a more precise estimate of the variant frequency in Thailand. It was detected in 6 (0.06%) individuals yielding an OR=45.2 (14.5-141.3) compared to the BrS cohort (p=5.6e-12). In addition to gnomADv3-EAS, the RE5 variant is also not detected in other genomic datasets of Asian ancestry (comprising a total of 75,948 individuals, Table S2), suggesting it is restricted to Thai or Southeast Asian populations.

None of the 9 BrS cases with the RE5 variant had rare or low frequency coding variants in *SCN5A*. When grouping cases by genotype, i.e. (1) RE5 variant, (2) ultra-rare *SCN5A* coding variants, (3) *SCN5A*:p.Arg965Cys and (4) no low frequency (gnomAD FAF<0.001) *SCN5A* coding variants, no statistically significant differences in clinical characteristics were observed (Table 1), although a significant leftward shift of ECG QRS axis was observed in cases with RE5 variant (7.88±52° vs 46.4±51°, p<0.05) with 2 cases displaying left QRS axis deviation (-30° to -150°).

However, cases with the RE5 variant were particularly symptomatic - 89% (8/9) had a history of sudden cardiac arrest (SCA), of which 7 had documented ventricular tachycardia/ventricular fibrillation (VT/VF), compared to only 58% of cases without any *SCN5A* variant. For one patient, analysis of their family pedigree revealed the variant largely segregated with phenotype, although one genotype-negative individual displayed a drug-induced BrS ECG pattern. However, this should be interpreted in the context of an appreciable false positive rate for this test^14^, and the fact that it is not atypical in BrS families for individuals lacking the familial pathogenic *SCN5A* variant to be phenotype-positive due to the complex genetic aetiology of the disease (Fig. 2A)^15^.

**Fig. 2.**
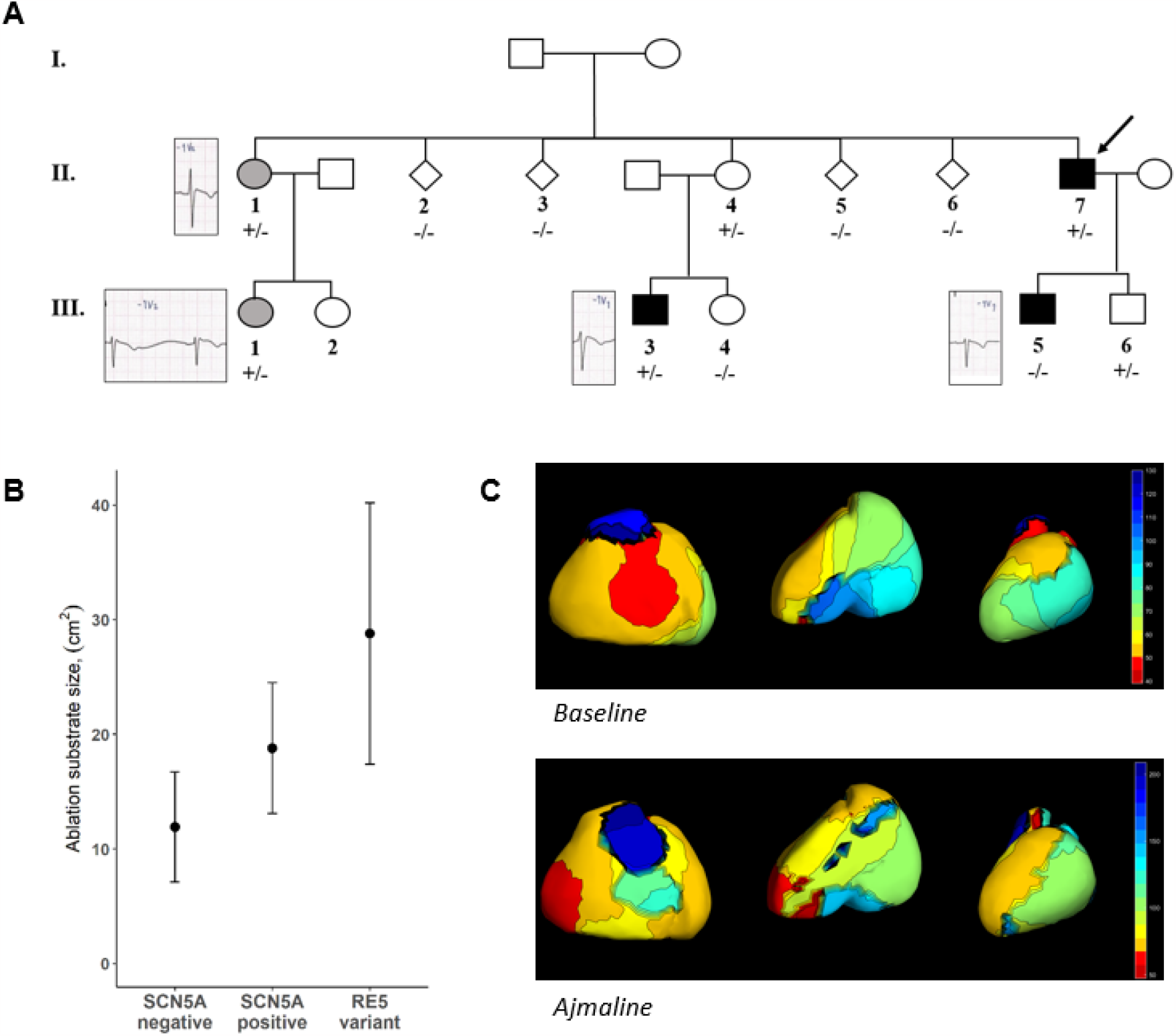
Clinical aspects of carriers of the RE5 variant. A) Family pedigree of one of the BrS patients carrying the RE5 variant (the proband is II-7 as indicated). One family member with the variant tested positive for BrS after ajmaline challenge (III-3, black fill) while two others had a borderline BrS marker on baseline ECG (II-1, III-1, grey fill). One individual also tested positive for BrS after ajmaline challenge despite not carrying the RE5 variant (III-5) – this is not atypical for SCN5A pathogenic variants in BrS families and reflects the complex genetic aetiology of the disease. No other non-carriers display the Brugada marker on the ECG. B) Epicardial substrate ablation areas (after ajmaline challenge) for BrS patients carrying the RE5 variant (n=3, substrate sizes of 40.0cm^2^, 29.2 cm^2^ and 17.2cm^2^), compared to data from Ciconte et al for carriers of pathogenic coding variants in SCN5A (SCN5A pos., n=49, mean/SD=18.8±5.7cm^2^) and BrS cases without SCN5A variants (SCN5A neg., n=146, mean/SD=11.9±4.8cm^2^)^17^. C) Baseline (above) and post ajmaline (below) epicardial mapping for the proband in the pedigree of part A.

Three RE5 patients underwent epicardial substrate ablation using the sodium channel blocker ajmaline to define the substrate areas of the right ventricular outflow tract and right ventricular inferior epicardium^16^. These procedures revealed large substrate areas more similar to *SCN5A*-positive than *SCN5A*-negative cases^17^, suggesting a substantial loss-of-function effect of the RE5 variant (Fig. 2B, 2C).

To assess the potential consequences of the RE5 variant, we performed a transcription factor (TF) motif scanning analysis using Homer^18^ on a 1,286 base pair stretch of the hs2177 enhancer sequence^19^ (which is fully encompassed by the RE5 region and includes the variant site), scanning for the presence of 1,847 motifs in the Homer^18^ and Jaspar^20^ databases. Nine variant-dependent motif recognition sites were detected; 8 exclusive to the wild type sequence and one to the RE5 variant sequence (Table S3). Of the 8 sites lost by the RE5 variant, 6 are different representations of the Mef2 motif (CTAWWWWTAG; -5 to +5)). The variant substitutes the A nucleotide at position +4 of the motif, previously shown to cause a strong reduction in binding affinity for Mef2^21^. This Mef2 binding sequence is conserved in evolution and its functionality is supported by Mef2 ChIP-seq data from mouse hearts^22^ (Fig 1D-E). The motif gained by the RE5 variant represents a binding site for the Gfi1B transcriptional repressor^23^.

In order to evaluate the functional consequences of the presence of the RE5 variant in the context of a human cardiomyocyte, we introduced the heterozygous variant into a human induced pluripotent stem cell (hiPSC) line. The PGP1 hiPSC line (details in Methods) was edited by the CRISPR/Cas9-mediated homology-directed repair method and two isogenic clones (A7 and C8) carrying the single nucleotide variant in heterozygous form were isolated and characterised (Fig. S2). HiPSC-CMs from the parental and heterozygous mutant lines were generated by adaptation of a protocol based on small molecules-mediated modulation of the cWnt-pathway^24^ (details in Methods).

We evaluated the potential cis-regulatory effect of the RE5 variant on *SCN5A* allelic expression through RNAseq analysis of hiPSC-CMs from the A7 variant line and the isogenic control (PGP1). To distinguish transcripts originating from the two alleles, we took advantage of the presence of a heterozygous SNP located in an exon adjacent to the RE5 intronic variant (rs7430407, GRCh38:3-38580976-T-C, located 596 bases from the RE5 variant) and determined their phase in the A7 line (details in Methods) (Fig. 3B). The allele-specific transcript output ratios in the hiPSC-CMs of the two isogenic lines (PGP1 and A7) were quantified and compared, with A7 hiPSC-CMs containing the RE5 variant displaying a 34% reduction of the *SCN5A* transcript allelic ratio compared to PGP1 controls (Z-test, p=0.0004) (Fig. 3C, Table S5). Of note, in mice, the two *Scn5a* alleles are independently regulated^6^. These data further indicate allele-specific reduced RE5 enhancer activity and *SCN5A* expression due to loss of the Mef2 motif as a consequence of the variant.

**Fig. 3.**
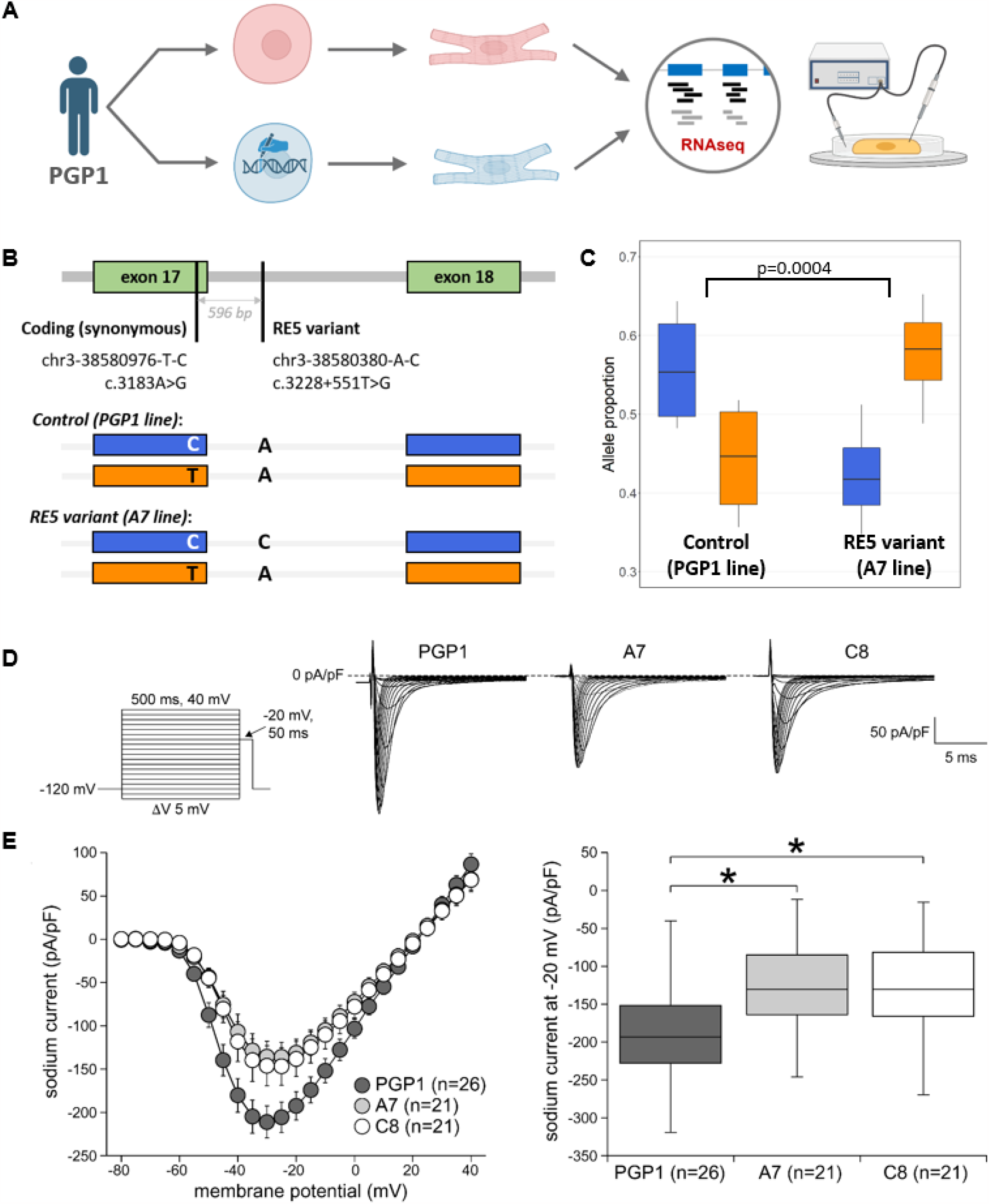
Experimental characterisation of the RE5 variant. **A**. Overview of experimental validation: Generation of human induced pluripotent stem cell derived cardiomyocytes (hiPSC-CMs) from PGP1 cell line – isogenic control (red) and RE5 variant line (blue), where the variant was introduced in the heterozygous state by CRISPR-Cas9 editing (graphic produced with Biorender). B. Allelic balance of SCN5A expression in the two lines was assessed using the heterozygous synonymous coding variant c.3183A>G present in the PGP1 line (596 bases upstream of the RE5 variant in exon 17), with the C exonic allele demonstrated to be in phase with the RE5 variant. C. Allelic balance of SCN5A expression in the control and RE5 variant lines as determined by RNAseq (4 biological replicates each) where p value refers to the Z-test testing differences in allelic expression ratios. D. Sodium current (I_Na_) characterization in control hiPSC-CMs (PGP1) and hiPSC-CMs of the two RE5 variant lines (A7 and C8), showing representative traces of I_Na_ activated during the first depolarizing pulses of the voltage clamp protocol shown in the inset. E. Average current-voltage (I-V) relationships of peak I_Na_ (left panel) and boxplots depicting I_Na_ densities (median and boxes represent interquartile range), determined at -20 mV (right panel). * indicates significance compared to CTRL (p<0.05; Kruskal-Wallis test, followed by Dunn’s comparisons).

I_Na_ measurements were performed in control hiPSC-CMs (PGP1) and in the hiPSC-CMs of the two isogenic lines (A7 and C8) with the RE5 variant. Fig. 3D shows typical I_Na_ recordings (−80 to 0 mV) from all groups and Fig. 3E (left) shows the average current-voltage (I-V) relationships of I_Na_. The I_Na_ density was significantly lower in hiPSC-CMs of both RE5 variant lines, with a 30% reduction at - 20mV (Fig. 3E, right). Neither the voltage dependency of activation (Fig. S3A) nor the voltage dependency of inactivation (Fig. S3B) were affected by the RE5 variant. The time constants of current inactivation, determined at a test potential of -20 mV, were not different in hiPSC-CMs of the two RE5 variant lines compared with the control (data not shown). In summary, the RE5 variant results in a decrease in I_Na_ density without changes in gating properties, concordant with the reduced *SCN5A* expression from the allele harbouring the RE5 variant.

In this study we have identified and validated a rare non-coding *SCN5A* enhancer variant associated with BrS in Thailand, one of the first examples of a rare regulatory variant of proven pathogenicity in a cardiac disease gene. The strong observed effect size of this variant (OR=20-45, 30% reduction in sodium current density) is reflected in the severe phenotype in BrS patients and may be explained by the predicted concurrent loss of a Mef2 motif (a critical family of cardiac transcription factors) and possible gain of a Gfi1 transcriptional repressor (although further studies will be needed to probe the exact mechanisms underlying *SCN5A* downregulation). This study highlights how detailed epigenetic knowledge across disease-relevant genomic loci can reduce the search space of the vast non-coding genome and address the challenge of missing heritability in rare disease.

The lower diagnostic yield of ultra-rare *SCN5A* coding variants in BrS patients from Southeast Asia compared to those of European ancestry is likely largely due to the increased prevalence of BrS in this region, i.e. the absolute rate of such variants is expected to be broadly similar across populations in the absence of specific founder variants (Fig. 4). Similarly, we previously described an 8.9% yield in Japanese BrS patients reflecting a disease prevalence intermediate between Europe and Southeast Asia^2^. The RE5 variant is found in approximately 1 in every 25 BrS patients from Thailand and highlights the potential of rare non-coding variation to account for missing heritability in diseases like BrS and, along with the low frequency *SCN5A*:p.Arg965Cys variant, partly explains the increased prevalence of this condition in this region (Fig. 4).

**Fig. 4.**
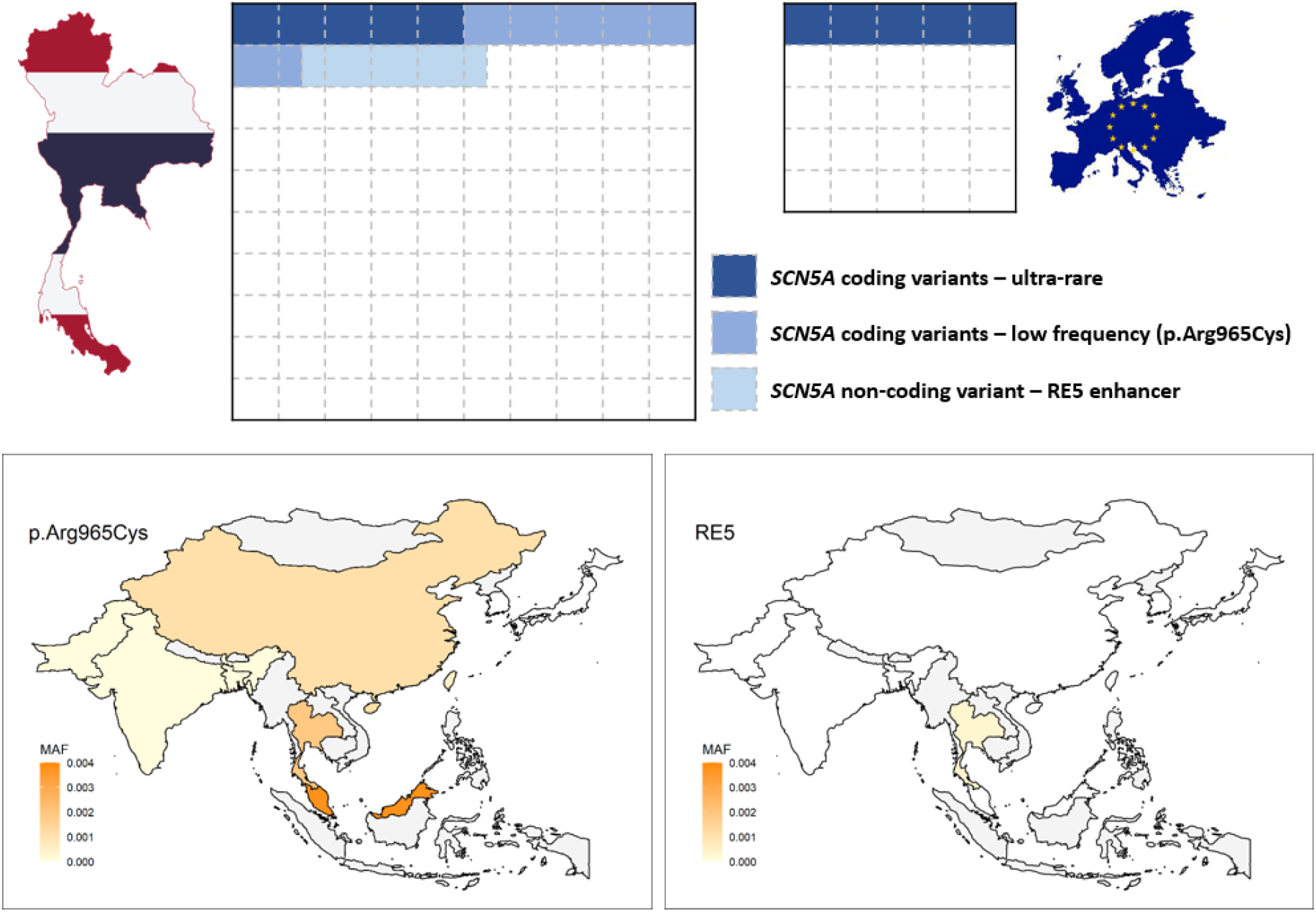
Top. Graphical representation of the genetic contribution of rare and low frequency variants to Brugada syndrome (BrS) in Thailand (left) and European ancestry populations (right), assuming an estimated four fold greater prevalence of BrS in Thailand. The absolute frequency of ultra-rare loss-of-function coding variants in SCN5A is likely to be broadly similar in both ancestries, but the diagnostic yield of such variants in BrS cohorts from Thailand is relatively lower (∼5% vs ∼20%) due to the different disease prevalence between ancestries. A proportion of the additional susceptibility in Thai BrS patients is explained by the low frequency SCN5A coding variant p.Arg965Cys (∼6.5%) and the non-coding RE5 enhancer variant described in this study (∼4%). **Bottom** – Population frequencies of SCN5A p.Arg965Cys and RE5 variants in South and East Asian countries – see Table S6 for details of datasets used and exact variant frequencies (grey = data not available).

The persistence of at least two recurrent *SCN5A* loss-of-function variants of intermediate to large effect size in the Thai population is intriguing given their association with sudden death as BrS risk factors. While selective pressure against pathogenic BrS variants may be relatively modest, as sudden death usually occurs in fourth or fifth decade, these variants (and likely other such risk factors in East Asian populations) appear to be restricted to this region. Several examples of disease risk factors that are protective against infectious diseases are known, such as the relationship between malaria and sickle cell trait or *APOL1* polymorphisms that protect against African sleeping sickness but increase the risk for chronic kidney disease^25^. Whether genetic variants associated with partially reduced sodium channel function confer similar protection is unknown, but would offer an intriguing hypothesis for the highly increased prevalence of BrS across Southeast Asia.

## Online Methods

### BrS and control cohorts

The BrS cases were recruited from seven participating institutions in Thailand. All patients were diagnosed with BrS according to the 2013 Heart Rhythm Society/European Heart Rhythm Association/Asia Pacific Heart Rhythm Society Expert Consensus Statement^26^. Blood samples were collected for DNA extraction and genotyping. The control samples were collected from volunteers who had no Brugada pattern (type 1) on standard 12-lead ECG. All cases and controls were of Thai ethnic origin by self-report. The study was approved by the Institutional Review Board of the Faculty of Medicine, Chulalongkorn University, Bangkok, Thailand (IRB No. 431/58). Informed consent was obtained from all participants. All methods were performed in accordance with relevant guidelines/regulations.

### Genome sequencing and analysis

#### Data generation and processing

The samples were sequenced by the Illumina FastTrack service on HiSeqX with a PCR-free library. Fastq files were aligned to the GRCh37 reference build using BWA Mem 0.7.12^27^ and converted to BAM using Samtools 1.3.1^28^. The data from the alignment and MarkIlluminaAdapters was merged with MergeBamAlignment and sorted with Sambamba 0.6.6^29^. Where a sample belonged to multiple read groups, the read groups were merged with sambamba merge. BAM files were split into chromosomes and each chromosome cleaned with Picard CleanSam, duplicates were removed with Sambamba markdup. Indels were realigned with GATK 3.5 RealignerTargetCreator and IndelRealigner. All chromosomes and non-chromosomes were merged back into one bam file with sambamba merge. Haplotypes were called with GATK HaplotypeCaller^30^. All GVCFs were merged into 10,000,000 base pair chunks with GATK3.8 CombineGVCFs and called with GenotypeGVCFs. All chunked VCFs were merged into chromosomes with GATK CombineVariants, and SNP and Indel variants were recalibrated with GATK VariantRecalibrator and ApplyRecalibration. The variants were annotated with SNPsift/SNPeff 4.1. The chromosomes were merged into one file with Picard. The pipeline was implemented in Snakemake^31^. Sample exclusion criteria were a missing genotype rate of >0.03, relatedness (PI_HAT>0.2, one of related sample group retained with priority for cases), high inbreeding rate (F>0.1) and sex check inconsistencies. Sample ancestry was determined using principal component analysis (PCA) with samples from the 1000 Genomes (1KG) dataset – all samples clustered with East Asian 1KG samples. A PCA of cases and controls was plotted to demonstrate ancestry matching.

#### Genetic variant analysis

All SCN5A coding variants were described with respect to the ENST00000333535 transcript. Coding variants were classified using the guidelines of the American College of Medical Genetics and Genomics and the Association for Molecular Pathology (ACMG-AMP)^32^ as previously described^2^. Non-coding variants at the *SCN5A-SCN10A* locus were assessed for enrichment in cases versus controls, using Fisher’s exact test (for samples with/without variants). The following regulatory regions at the locus were queried for variants (all coordinates based on GRCh38): RE1/*SCN10A* enhancer (chr3:38725264-38728203); RE2/*SCN10A* enhancer (chr3:38721868-38725264); *SCN10A*-short promoter (chr3:38716810-38719605); *SCN5A* promoter (chr3:38642253-38652554); RE4/*SCN5A* enhancer (chr3:38613169-38618856); RE5/*SCN5A* enhancer (chr3:38575835-38584663); RE6-9/*SCN5A* super enhancer (chr3:38526650-38546621), as well as the intermediate sequence between these regions (Fig. 1A)^6^. To assess for linkage disequilibrium between variants, r^2^ values were calculated for each variant pair in this region (with a combined minimum case and control allele count of 3). Variants of interest were further annotated with CADD and FINSURF scores^13^, frequency in gnomADv3 East Asian genome sequences (n=2,604) and conservation of affected base across species using Ensembl v109 (Feb 2023). The frequency of the putative causal variant was also assessed in other genomic databases with individuals of Asian ancestry – the Genomics Thailand database of 10,043 individuals of Thai ancestry, the Singaporean SG10K_Health v5.3 database of individuals of Chinese, Indian and Malay ancestry^33^, the jMORP/54KJPN database of individuals of Japanese ancestry^34^ and the KOVA database of individuals of Korean ancestry^35^.

#### Clinical characteristic statistical analysis

Clinical characteristics including demographic data, history of cardiac events and electrocardiogram (ECG) parameters were compared among the four genotype groups (RE5 variant, ultra-rare *SCN5A* coding variants, *SCN5A*:p.Arg965Cys and no low frequency *SCN5A* coding variants). Statistical analyses were performed using chi-square or fisher exact tests for categorical variables and ANOVA for continuous variables.

#### Transcription factor motif analysis

Transcription factor (TF) motif scanning analysis was performed using Homer^18^ to detect the presence of 1,847 motifs in the Homer^18^ and Jaspar^20^ databases for the wild type and variant sequences in the enhancer region of interest. The enhancer region was further characterised using human and mouse ChipSeq datasets (see Table S4 for details).

### Generation of hiPSC-CMs

#### HiPSCs culture, genome editing and characterisation of clones

The Personal Genome Project 1 hiPSC line, PGP1-SV1, referred to as PGP1 in this manuscript (detailed information at https://arep.med.harvard.edu/gmc/PGP_cells.html), as well as the derived edited clones were maintained on growth factor-reduced Matrigel Matrix (Corning)-coated plates in presence of mTeSR1 medium (Stemcell Technologies). Cells were passaged every 3-4 days using 0.5mM EDTA (Life Technologies) in PBS without CaCl_2_ and MgCl_2_ and re-plated in presence of Rho kinase inhibitor (Fasudil HCl, Selleck Chemicals) for the first 24 hours. Cells were maintained at 37°C, with 5% CO_2_ and 20% O_2_. The RE5 GRCh38:chr3-38580380-A-C variant was introduced in heterozygous state in the PGP1 hiPSC line by CRISPR/Cas9-mediated editing (Synthego). The Cas9 and gRNA, as ribonucleoprotein, and the single strand oligodeoxynucleotide (ssODN) carrying the single nucleotide variant for homology-directed repair (sequences in Table S7) were introduced by nucleofection. Two heterozygous, edited clones (A7 and C8) were selected after Sanger sequencing of their genomic DNA (primers in Table S7, Fig. S2A). Quality controls were performed on these edited clones which confirmed karyotype integrity by KaryoStat Assay (Fig. S2B) and pluripotency by PluriTest Assay (Fig. S2C) (Synthego). One predicted off-target genomic locus, exhibiting two mismatches with the guideRNA spacer sequence, was identified by the Wellcome Sanger Institute Genome Editing (WGE) off-target finding tool (https://wge.stemcell.sanger.ac.uk/find_off_targets_by_seq) at position GRCh38:chr8-15043569-15043591. A ∼200bp window of genomic DNA from the A7 and C8 clones, centred around this predicted potential off-target site, was sequenced by Sanger sequencing to detect any sequence alterations (primer sequences in Table S7). No sequence alterations were detected in in the two edited clones (A7 and C8).

#### hiPSC-CMs generation

The parental PGP1 and edited A7 and C8 hiPSC lines were maintained in undifferentiated state in presence of mTeRS1 medium on Matrigel Matrix-coated plates until they reached 65-80% confluence. Differentiation was then performed as described by Maas *et al*.^24^ without cardiomyocytes expansion and with the following modifications. Differentiation was started in RPMI 1640 medium containing 2% B27 supplement without insulin (both by Gibco/Thermo Fisher Scientific) (hereafter indicated as RPMI/B27−) and 6 µM CHIR99021 (Selleck Chemicals). After 24 hours a volume of RPMI/B27− medium without CHIR99021 equal to 2/3 of the original differentiation medium was added. On day 3 of differentiation 1/3 of the original volume of RPMI/B27− medium was further added to gradually dilute the CHIR99021 concentration. The differentiation protocol continued with 3-days treatment with 2 µM Wnt-C59 (MedChemExpress; Wnt signaling pathway inhibitor) in RPMI/B27− medium, without medium change. After Wnt signaling inhibition the culture was maintained until day 43 in RPMI/B27− medium alone, with medium change every 3-4 days. On day 44, a 7-8 days metabolic-selection that enriches the culture for cardiomyocytes was started by switching the medium to RPMI 1640 without glucose (Gibco/ThermoFisher Scientific) supplemented with 500 µg/mL bovine serum albumin (Sigma-Aldrich/Merck) and 8 mM Na-L-lactate (Sigma-Aldrich/Merck) with medium change every other day (adapted from Tohyama *et al*.^36^). The culture was then switched to RPMI 1640/B27− for 1 day before proceeding to either perform RNA isolation or to dissociate the cells for electrophysiological analysis.

### RNA sequencing

#### RNA isolation

Total RNA was isolated from hiPSC-CMs derived from 4 independent batches of differentiation of the parental PGP1 and heterozygous RE5 variant-carrying A7 hiPSC isogenic lines. Cardiomyocytes cultures were lysed by TRIzol Reagent (Ambion/Life Technologies) followed by chloroform addition (200ul/ml of TRIzol Reagent) and hydrophilic phase isolation after centrifugation at 4°C. RNA was then precipitated in presence of isopropanol, pelleted by centrifugation at 4°C and washed twice with 75% ethanol. The RNA pellet was then briefly air-dried and resuspended in RNAse-free water.

#### Determination of phase between the RE5 variant and an exonic SNP

To determine the SCN5A transcript output from each allele of the A7 and PGP1 lines using bulk messenger RNA sequence analysis, we took advantage of the presence of a heterozygous SNP (GRCh38:3-38580976-T-C; rs7430407) in *SCN5A* exon 17. The phase between the SNP and the RE5 variant in the A7 line was determined by amplifying an 802 base pair region encompassing the 596bp interval between the rs7430407 SNP and the RE5 variant (primers in Table S7), followed by cloning and Sanger sequencing. This demonstrated that the C-allele of the RE5 variant was in cis with the C-allele of rs7430407 (see cartoon in Fig. 3B).

#### Bulk RNA sequencing and analysis

Quality and integrity of the isolated RNA was determined by 4200 TapeStation (Agilent) analysis which indicated a RIN score above 9 for all samples. Cardiomyocytes from the parental PGP1 and isogenic A7 hiPSC lines were used for bulk RNA sequencing. Library preps were made after ribosomal RNA depletion by KAPA Total RNA HyperPrep with RiboErase kit (Roche). Sequencing was performed on an Illumina platform NovaSeq S4.300 in pair-end mode with read length of 150bp (PE150) and a sequencing depth of 60 million reads per sample.

We used STAR^37^ to map reads to the hg19 build of the human genome/transcriptome. We used VarScan 2^38^ to detect variants and their allele specific occurrence within the samples. We used a minimum base quality Phred score of 15, a minimum read depth of 10 and a minimum variant allele frequency threshold of 0.20 in VarScan 2 for variant detection. To determine whether a variant was specifically enriched or depleted in either cell line, indicative for allele-specific enhancer activity, a Z-test was performed on the observed allele frequencies (p<0.05; Z-test)^39^.

### Electrophysiology patch clamp analysis

#### Preparation of hiPSC-CMs for single cell electrophysiological analysis

The hiPSC-CMs cultures were dissociated to obtain single cardiomyocytes as follows. The culture was incubated at room temperature for 7-10 minutes in a mixture (50/50) of Hanks Balanced Salt Solution (HBSS, Gibco/Thermo Fisher Scientific) without CaCl_2_ and MgCl_2_ and HBSS with CaCl_2_ and MgCl_2_. After HBSS mixture removal the culture was incubated for 15-20 minutes at 37°C in presence of TrypLE Select Enzyme 10x (Gibco/Thermo Fisher Scientific). The cells were then detached from the surface by pipetting, the enzyme solution was diluted with addition of RPMI/B27− medium and the cells in suspension were collected by centrifugation. The cell pellet was further dissociated in presence of Liberase TM Research Grade (Roche/Merck) as previously reported^40^. The hiPSC-CMs, seeded on Matrigel Matrix-coated cover glasses (VWR International GmbH) in RPMI 1640/B27− medium, were analysed 8-10 days after dissociation.

#### Data Acquisition

I_Na_ was recorded from hiPSC-CMs using an Axopatch 200B amplifier (Molecular Devices, Sunnyvale, CA, USA). Voltage control, data acquisition, and analysis were realized with custom software^41^. Signals were low-pass-filtered with a cutoff of 5 kHz and digitized at 20 kHz. Cell membrane capacitance (C_m_) was calculated by dividing the time constant of the decay of the capacitive transient after a −5 mV voltage step from −40 mV by the series resistance. Series resistance was compensated for by at least 80%. Following the procedure of Veerman et al.^42^, we selected single, spontaneously beating hiPSC-CM_S_ showing regular, synchronous contractions in modified Tyrode’s solution, after which this extracellular solution was switched to a solution suitable for specific I_Na_ measurements. Modified Tyrode’s solution contained (in mmol/L): NaCl 140, KCl 5.4, CaCl_2_ 1.8, MgCl_2_ 1.0, glucose 5.5, HEPES 5.0; pH 7.4 (NaOH).

#### Sodium current measurements

I_Na_ was recorded at room temperature (∼20°C) using the ruptured patch-clamp technique. Extracellular solution contained (in mmol/L): NaCl 20, CsCl 120, CaCl_2_ 1.8, MgCl_2_ 1.0, glucose 5.5, HEPES 5.0; pH 7.4 (CsOH). Nifedipine (10 µmol/L; Sigma) was added to block the L-type Ca^2+^ current in hiPSC-CMs^43^. Patch pipettes were pulled from borosilicate glass (GC100F-10; Harvard apparatus, UK) and had resistances of ∼2.5 MΩ after filling with the solution consistent of (in mmol/L): NaCl 3.0, CsCl 133, MgCl_2_ 2.0, Na_2_ATP 2.0, TEACl 2.0, EGTA 10, HEPES 5.0; pH 7.2 (CsOH). I_Na_ was measured using a double pulse protocol from a holding potential of -120 mV (see inset Fig. 3D; cycle length of 5 seconds). The first pulse was used to determine the I-V relationship and voltage dependency of activation; the second pulse was used to analyze the voltage dependency of inactivation. I_Na_ was defined as the difference between peak and steady state current and current densities were calculated by dividing current amplitude by C_m_. Voltage dependence of activation and inactivation curves were fitted with Boltzmann function (I/I_max_=A/{1.0+exp[(V_1/2_ −V)/k]}), where V_1/2_ is the half-maximal voltage of (in)activation and k, the slope factor (in mV). The time course of current inactivation was fitted by a double-exponential equation: I/I_max_=A_f_ ×exp(-t/τ_f_)+A_s_×exp(-t/τ_s_), where A_f_ and A_s_ are the fractions of the fast and slow inactivation components, and τ_f_ and τ_s_ are the time constants of the fast and slow inactivating components, respectively.

#### Statistics

Data are expressed as mean±SEM, unless stated otherwise. Statistical analysis was carried out with SigmaStat 3.5 software (Systat Software, Inc., San Jose, CA, USA). Normality and equal variance assumptions were tested with the Kolmogorov–Smirnov and the Levene median test, respectively. Significance between parameters was tested using one-way repeated measures ANOVA followed by pairwise comparison using the Student–Newman–Keuls test or by the Kruskal-Wallis test followed by pairwise comparisons with Dunn’s method. P<0.05 defines statistical significance.

## Data Availability

All data produced in the present work are contained in the manuscript.

## Supplementary Figures

**Fig. S1.**
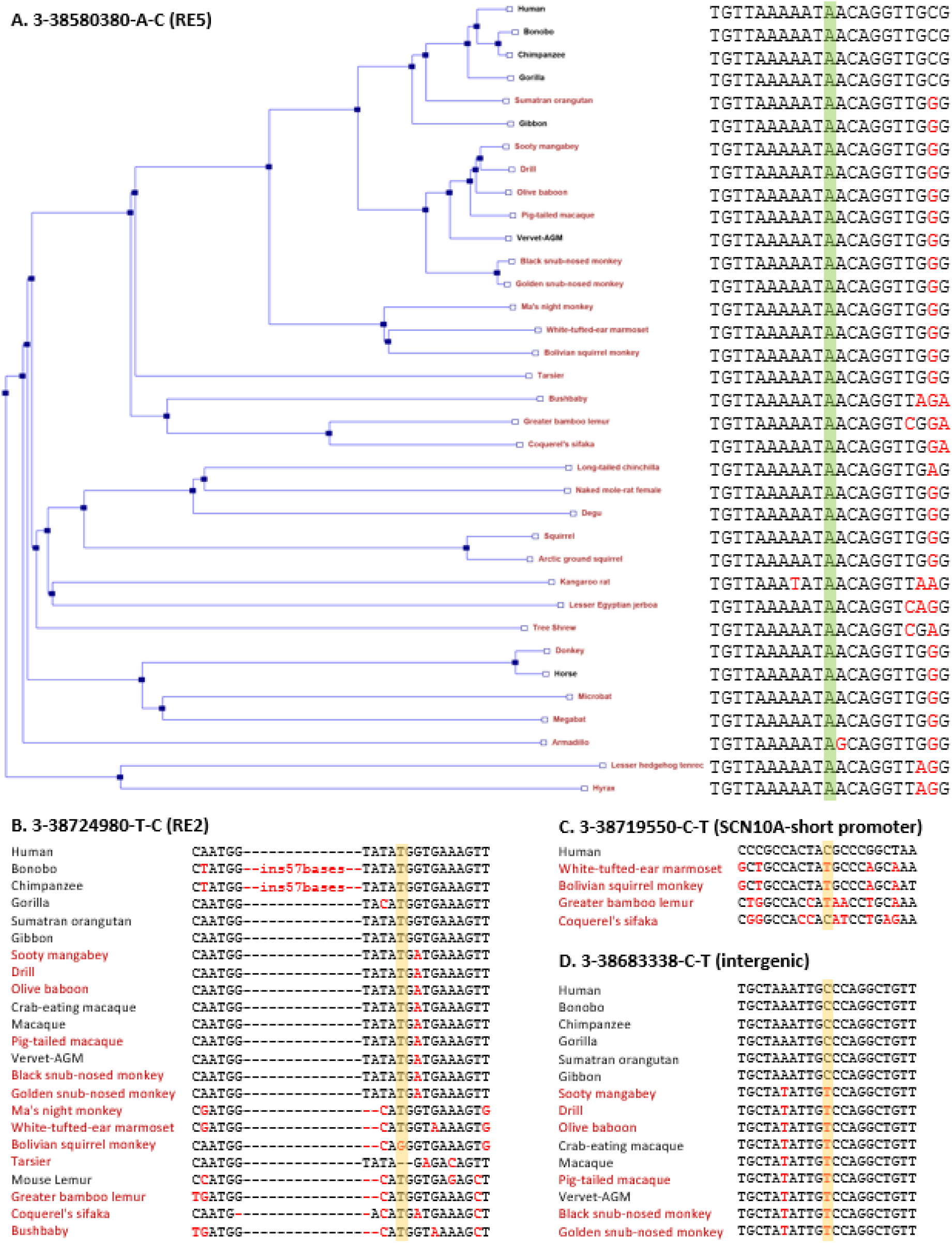
Conservation across species for the affected bases of four candidate non-coding variants at the SCN5A/SCN10A locus. Data is from Ensembl version v109 (Feb 2023), species with high quality assemblies are labelled in black and species with low quality assemblies in red. For each variant, species with orthologous alignments for the affected base and 10 flanking 5’ and 3’ bases are displayed. **A**. For the RE5 3-38580380-A-C variant, all eutherian mammals are aligned (55/90 species with no alignment), with complete conservation across the variant base (shaded in green). **B**. For the RE2 3-38724980-T-C variant, the variant base (shaded in orange) is not fully conserved across primates (0/23 species with no alignment), with several insertion/deletion gaps in close proximity. **C**. For the SCN10A-short promoter 3-38719550-C-T variant, the variant base (shaded in orange) is not conserved across primates (18/23 species with no alignment). **D**. For the intergenic 3-38683338-C-T variant, the variant base (shaded in orange) is not conserved across primates (8/23 species with no alignment).

**Fig. S2.**
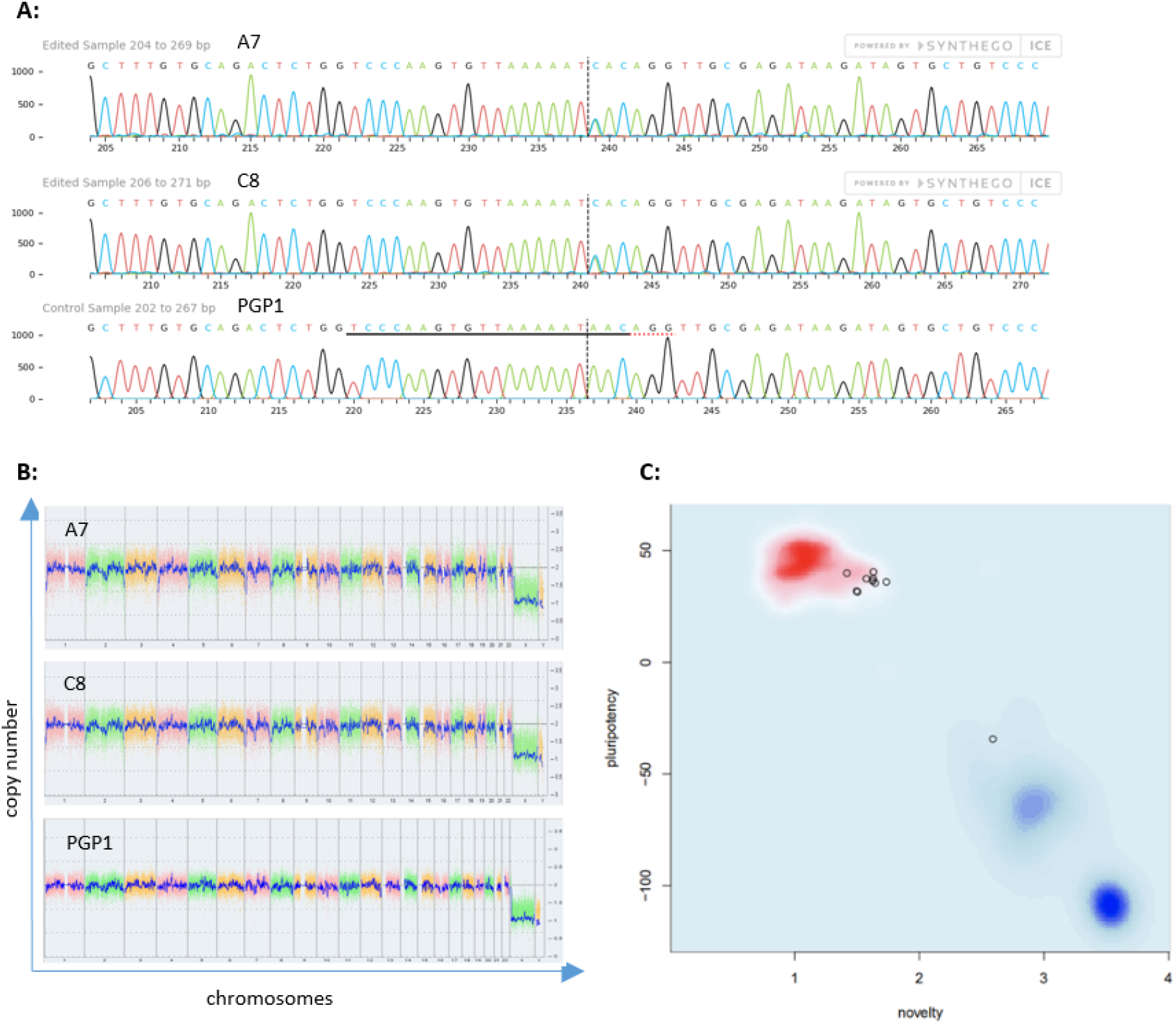
Quality control data on the edited hiPSC clones A7 and C8 carrying the heterozygous chr3-38580380-A-C variant. **A.** Sanger sequencing chromatograms of edited and parental (PGP1) hiPSCs showing the introduction of the heterozygous A/C SNP in the A7 and C8 clones. **B**. Karyotype analysis by KaryoStat Assay (Applied Biosystems) indicated that genomic integrity is maintained in the edited clones. **C**. A Pluritest Assay (Thermo Scientific) performed on the edited A7 and C8 clones (bold circles) showed the similarity of the edited clones transcriptome to the reference set of >450 samples (red).

**Fig. S3.**
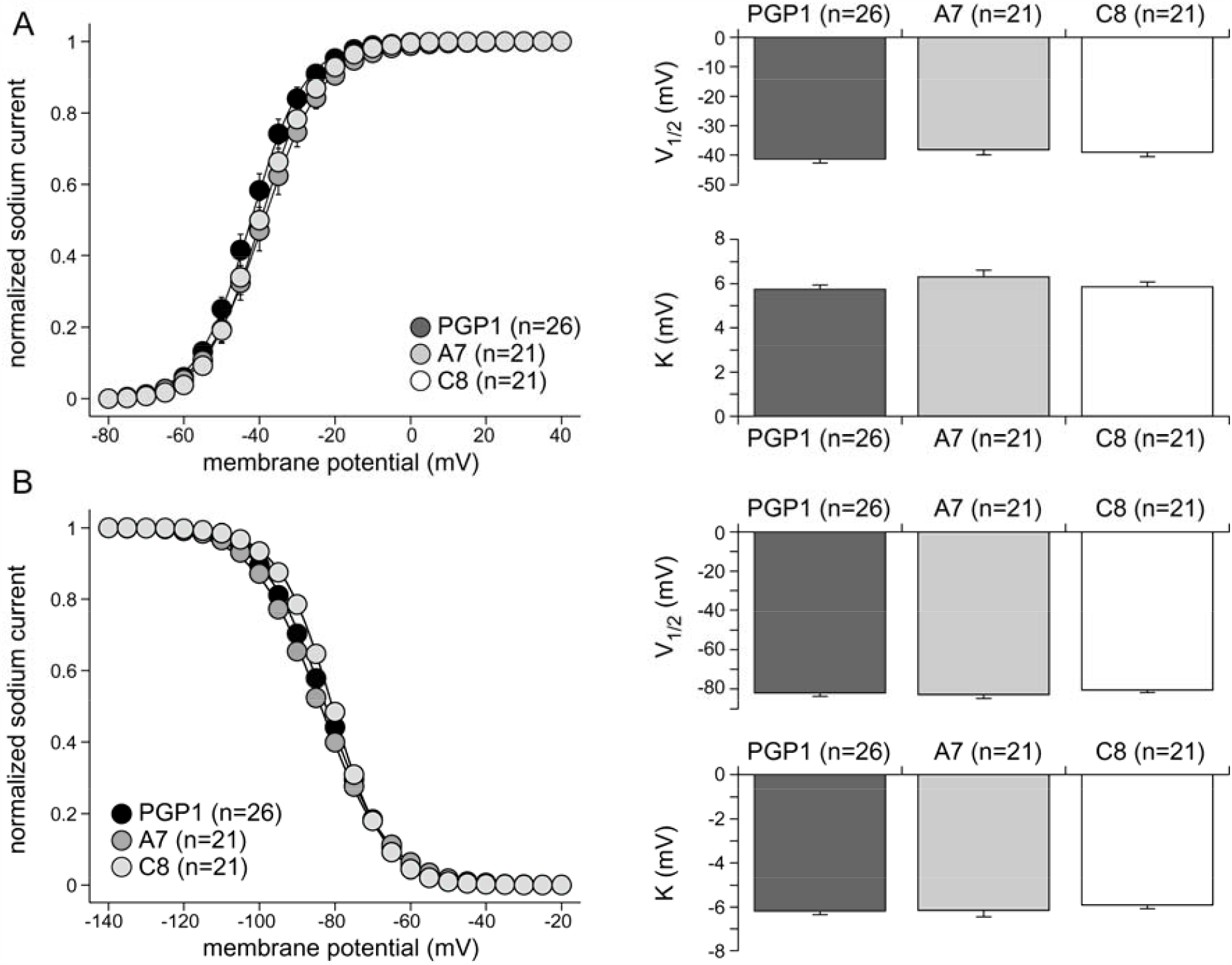
Sodium current (I_Na_) characterization in control hiPSC-CMs (PGP1) and in hiPSC-CMs of the two RE5 variant lines (A7 and C8). **A.** Average voltage dependence of activation curves (**left panel**) and average V_1/2_ and k values (**right panels**). Solid lines represents the fitted Boltzmann function (I/I_max_=A/{1.0+exp[(V_1/2_ −V)/k]}). B. Average voltage dependence of inactivation curves (**left panel**) and average V_1/2_ and k values (**right panels**). Solid lines represents the fitted Boltzmann function (I/I_max_=A/{1.0+exp[(V_1/2_ −V)/k]}).

## Acknowledgments

We acknowledge the National Biobank of Thailand, National Science and Technology Development Agency for their assistance in gathering variant population frequency from the Genomics Thailand preliminary data. J.M. received support by the Second Century Fund (C2F), Chulalongkorn University. Y.P., K.N. and A.K. received support from the National Research Council of Thailand. C.W. received support from the Yin Shu-Tien Foundation, Taipei Veterans General Hospital-National Yang-Ming University Excellent Physician Scientists Cultivation Program (No.108-V-A-013). E.R.B. received support from the Robert Lancaster Memorial Fund and UKRI. V.M.C received support from EIC Pathfinder Challenges (Nav1.5-CARED) and the Netherlands Organization for Scientific Research (OCENW.GROOT.2019.029). C.R.B. received support from the Dutch Heart Foundation (CVON PREDICT2), the Netherlands Organization for Scientific Research (VICI fellowship, 016.150.610), Fondation Leducq (17CVD02) and EIC Pathfinder Challenges (Nav1.5-CARED).

